# Using RT-PCR Testing to Assess the Effectiveness of Outbreak Control Efforts in São Paulo State, the Pandemic’s Epicenter in Brazil, according to Socioeconomic Vulnerabilities

**DOI:** 10.1101/2020.10.29.20221960

**Authors:** Tatiane C M Sousa, Natália P Moreira, Jose E. Krieger, Isabel S C Rosa, Marcela M. Zamudio, Maria Amélia S M Veras, Brigina Kemp, Lorena G. Barberia

## Abstract

**Background:** The testing of infected persons with SARS-CoV-2 is one of the cornerstones of an effective strategy deployed for pandemic control. The public health diagnostic effort is particularly important in regions with a critical transmission scenario and in vulnerable populations in these districts, such as São Paulo state, the Brazilian epicenter of the COVID-19 pandemic.

**Methods:** We developed an RT-PCR testing intensity effort index (RT-PCR TIEI) composed of seven indicators to assess the survelliance efforts in the São Paulo State. We used dynamic time-series cross-sectional models to analyze the association between the RT-PCR TIEI, the population living under high socioeconomic vulnerability levels, dependent on public health service (SUS), per capita income, and population density.

**Results:** On average, the RT-PCR TIEI score was 21.07. In the long-run, the RT-PCR TIEI is negatively associated with socioeconomic vulnerability (p-value=0.000, 95% CI −0.887, - 0.811), with a higher proportion of the population dependent on SUS (p-value= 0.000, 95% CI −0.871, −0.805), per capita income (p-value= 0.000, 95% CI −0.849,-0.792) and with population density (p-value=0.000, 95% CI −0.853; −0.801).

**Conclusion:** Testing efforts declined as the pandemic advanced, and the the lowest RT-PCR TIEI values were found in the most socioeconomic vulnerable RHDs. Local public laboratory presence was a predictor of a higher score. Thus, the low testing RT-PCR efforts and local laboratory inequalities affected surveillance capability, especially for socioeconomic vulnerable populations.

## Introduction

Governments across the world have demonstrated that more intensive efforts to identify infected individuals are essential to an effective containment of COVID-19 along with other non-pharmaceutical interventions (1) (2). Molecular tests, commonly referred to as RT-PCR (Reverse Transcription-Polymerase Chain Reaction) tests, are considered the gold standard as they detect the virus’s RNA in infected individuals and who are potentially transmitting the virus to other individuals. In turn, serological tests identify antibodies produced following a SARS-CoV-2 infection (3). Due to these differences, RT-PCR tests are considered the first-best test for controlling the transmission of SARS-CoV-2 and an essential part of a surveillance strategy that should also include identifying and isolating infected individuals and ensuring the quarantine of people who were in contact with those who tested positive (4). In this study, we developed a new index to measure and evaluate the intensity effort of RT-PCR testing within the public health system based on the best-recommended guidelines in testing effort, especially those recommendations issued by the World Health Organization (WHO) (4) (5) (6) and the Centers for Disease Control and Prevention (CDC) (3) (8).

We then analyzed the proposed index to assess the RT-PCR testing intensity efforts undertaken by São Paulo and its 17 Regional Health Departments (RHD), considering the public health system. São Paulo state, one of the most populous states in Brazil (9), has been an important epicenter of the country’s pandemic, and has 62% of its population depending exclusively on the public health system. There has been variation in the RT-PCR testing efforts across the state’s regions over the past six months, and since April 2020, the labs in the public health system are centrally managed by São Paulo’s state government (10).

Initially, in late February and March 2020, the earliest cases were registered in the capital and few other cities outside the Metropolitan Region (RHD I), but the cases were largely concentrated in the São Paulo metropolis (11). Since June 2020, the state’s interior has surpassed the capital in the number of COVID-19 registered cases (12). Simultaneously, in June, the state loosened non-pharmaceutical interventions contingent on assessing the pandemic’s evolution in each RHD (13). As of August 31, 2020, the capital accounted for 32% of the cases, and the rest of the state accounted for 68% of the total cases.

## Methods

### Study design and participants

We developed an RT-PCR testing intensity effort index (RT-PCR TIEI) and conducted an observational time-series cross-sectional study to apply the proposed index to assess the RT-PCR public testing efforts in São Paulo state and its 17 Regional Health Departments (RHD) (Appendix 1). Given time lags in data reporting, we analyzed information reported from epidemiological week 10 to week 35 (March 1st to August 29, 2020).

### Data Sources

The official data for the state of São Paulo is based on several government sources. Specimens of anonymized daily data on the RT-PCR tests conducted in all public laboratories and the number of daily new confirmed cases in the state of São Paulo, including mild and severe cases, are drawn from the Health Secretariat’s Intelligent Monitoring System (SIMI) (12). For testing, this data source provides the number of RT-PCR performed in the network of public labs for hospitalized and ambulatory patients.

Data on the 2020 population and social vulnerability were provided by the Foundation for Statewide System Data Analysis (14) (15). The 2010 Social Vulnerability Index of São Paulo State is based on five socioeconomic and four demographic indicators (15). Social vulnerability is measured as the percentage of people living in each of the 17 RHD under high or very high social vulnerability (including urban and rural areas). For each RHD, these values represent the mean value of all municipalities in the specific region.

We also obtained the proportion of the population which was exclusively dependent on the Brazilian public health system, known as Sistema Único de Saúde (SUS) in Portuguese, in May 2020. These data are provided by the Information Health System of the Secretariat of São Paulo State (16). The per capita income estimates are based on 2010 census data (17).

### Index of Intensity Effort of RT-PCR Testing Composition

To identify specific policies that should be included to develop an RT-PCR Testing Intensity Effort Index (RT-PCR TIEI), we analyzed WHO and CDC technical guidelines and daily press conferences that specifically addressed testing (3) (4) (5) (6) (8), and other academic publications and platforms (7) (18) (19) (20). Although the CDC is a U.S. institution, its recommendations can be applied in Brazil considering its expertise and experience in surveillance strategies and the COVID-19 pandemic similarities in Brazil and the USA, with stabilization of the number of cases and deaths at a high level (21).

The RT-PCR TIEI is based on seven indicators, as shown in Table 1, the composite score is an additive index re-scaled to range from zero (lowest) to 100 (highest level of testing effort). The first five indicators are based on testing surveillance policies per se; one indicator includes a measure to assess if testing is being used for contact tracing, and the last indicator aims to evaluate the quality of the information provided from official sources. The maximum score is attributable to the observance of mainly WHO and CDC recommendations, and the remaining scores are declining ordinal measures with a minimum value of zero.

**Table 1.**
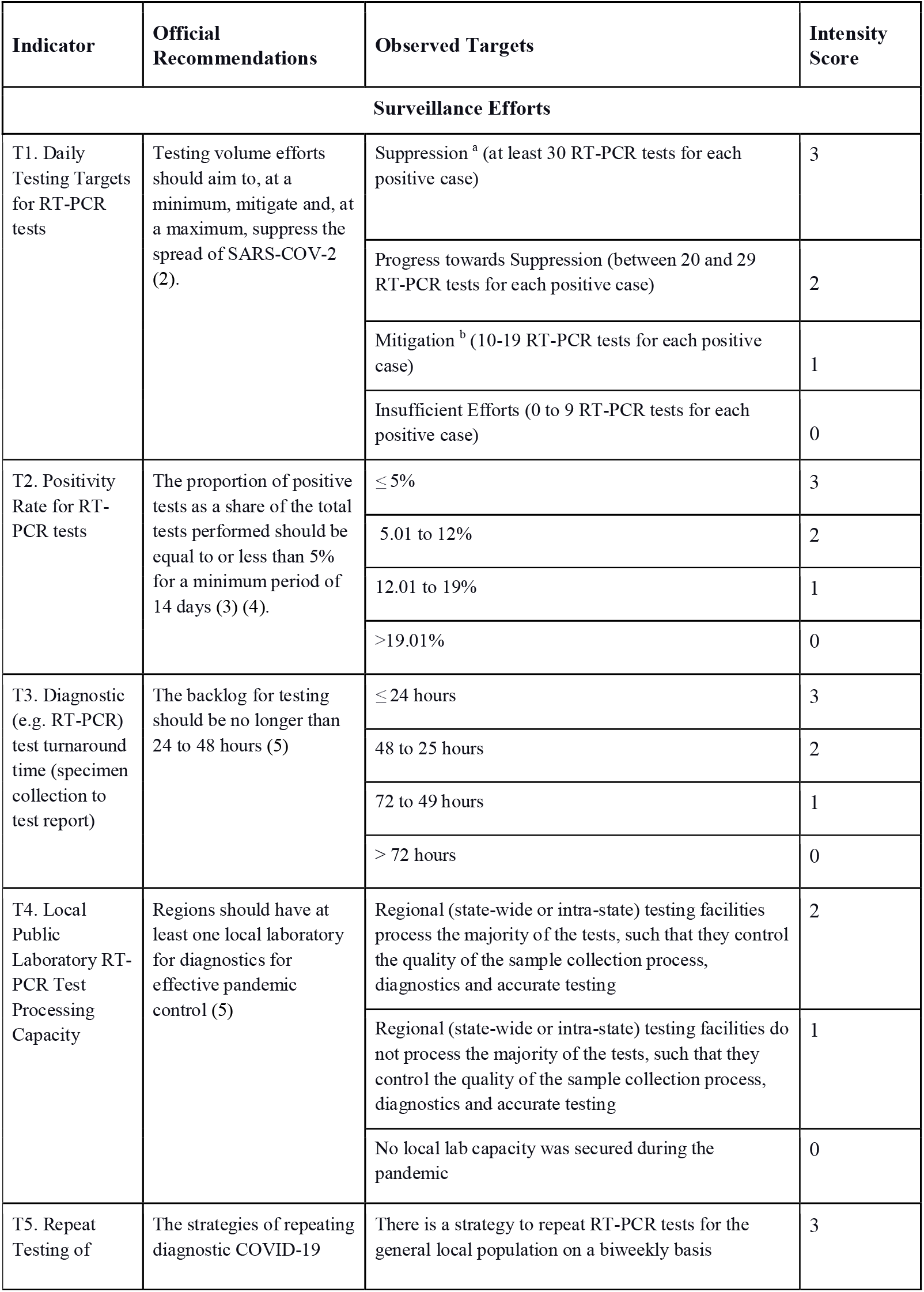
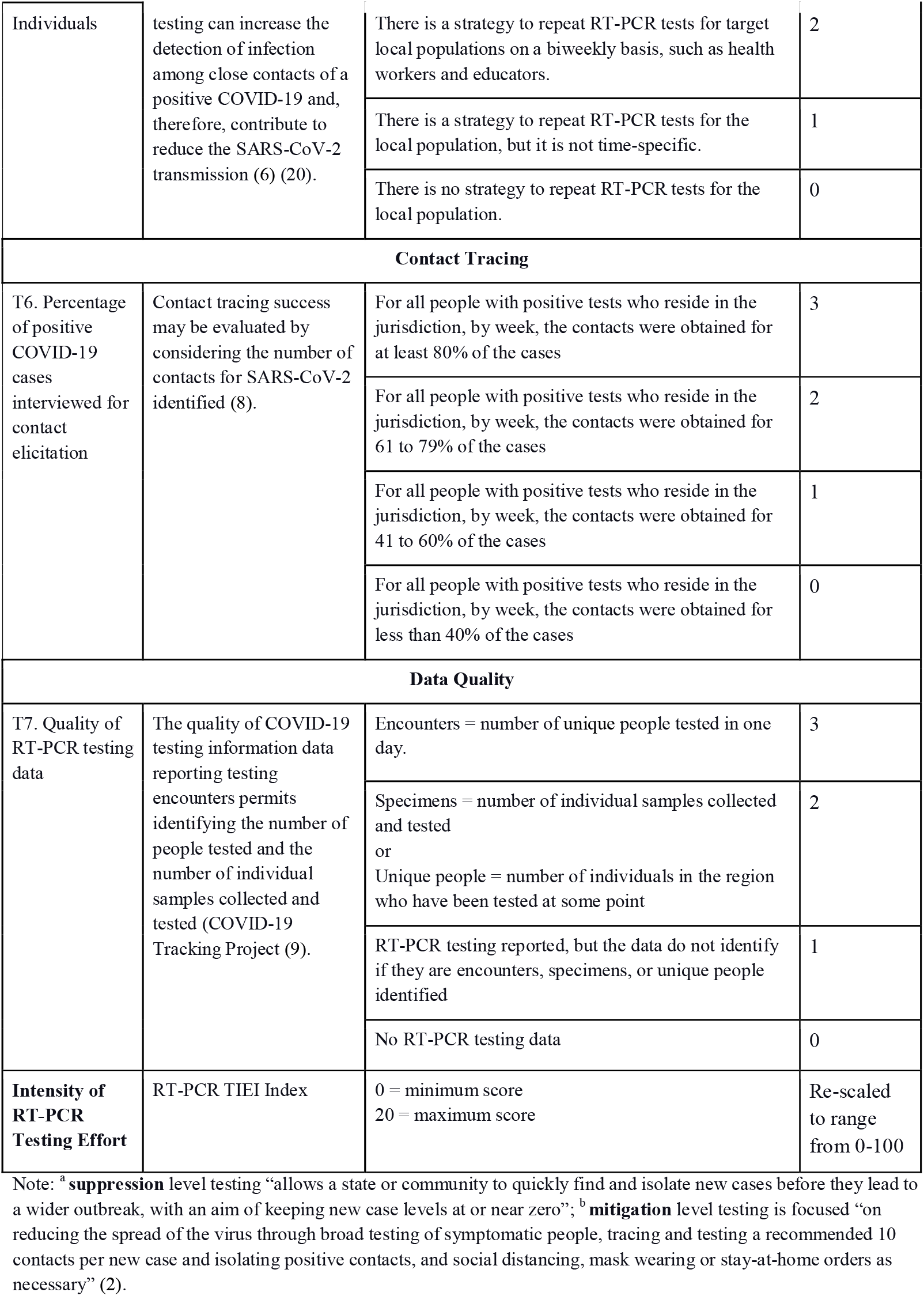
The Seven Indicators of the RT-PCR Testing Intensity Effort Index (RT-PCR TIEI).

### Statistical analysis

We estimated multivariate regression models to examine the association between the RT-PCR TIEI scores (dependent variable), per capita income, population density, the proportion of the population that depends on the SUS and living under high social vulnerability across 17 RHDs in São Paulo State in epidemiological weeks 10 to 35. The regression models included an intercept, and the lag of the RT-PCR TIEI and standard errors were clustered by RHD (22). In our analyses, the p-values less than 0.05 were considered statistically significant. All analyses were performed using STATA version 16.

## Results

As of August 31, 2020, the state of São Paulo had performed 3,049,073 tests, which is the highest number of RT-PCR tests of any state in Brazil, or 6,640 tests performed per 100,000 people (11). All RHD received the highest score for T1 (=3) in the first weeks analyzed (March). During that period, there were at least 30 RT-PCR tests for every positive case. After this period, all RHDs received the lowest possible score for T1 (=0) for the majority of the weeks analyzed. In these cases, the ratio of RT-PCR tests and COVID-19 confirmed cases between 0 and 9 implies that for each registered COVID-19 case, there were less than 10 RT-PCR tests performed in a week. Only three RHDs received a score of 2 for T1, but these scores were never maintained for more than one week in any RHD. In those cases, there were at least ten RT-PCR tests conducted for each positive case in a given week (generally in weeks 12 and 14).

**Figure 1.**
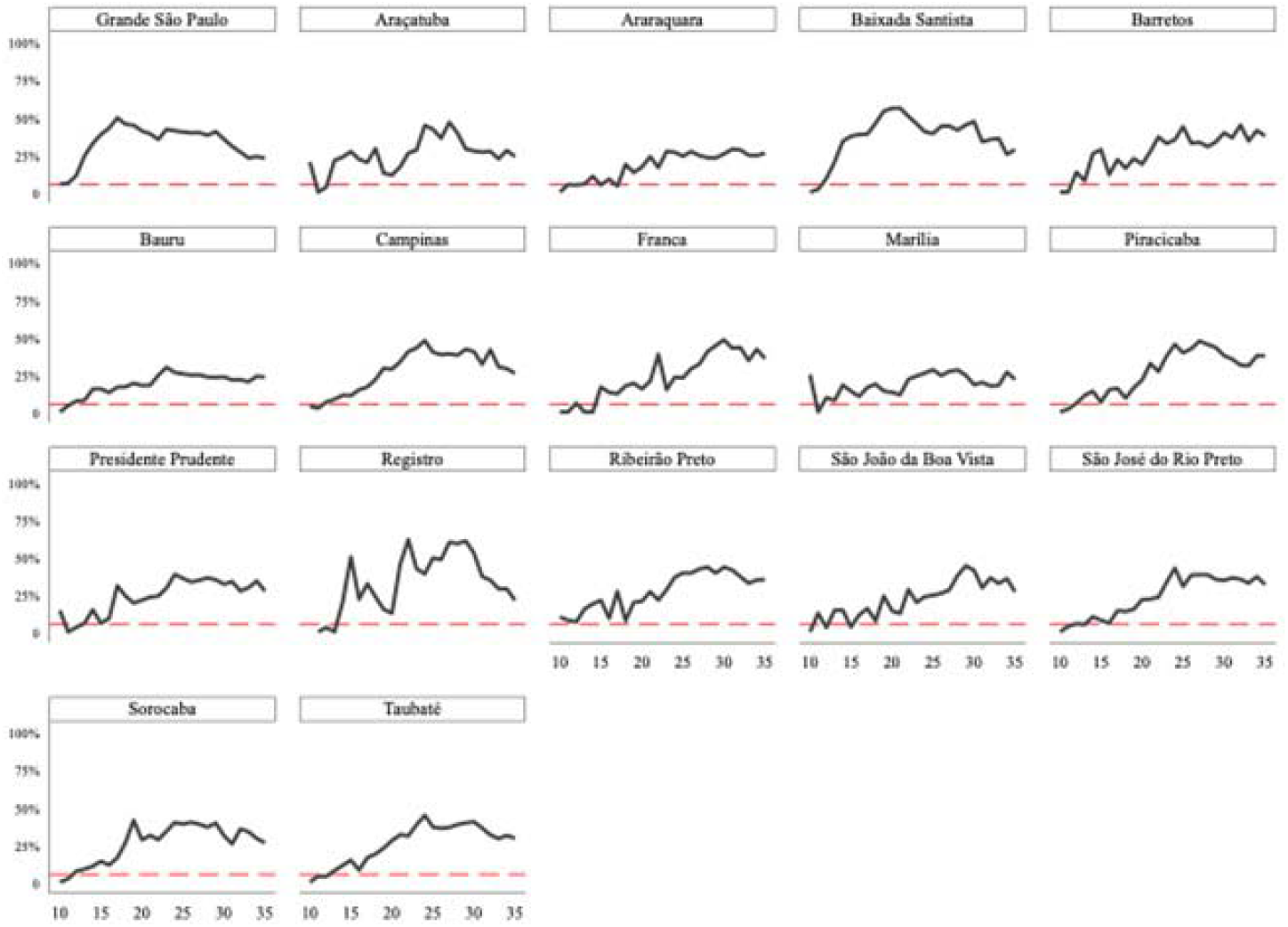
The RT-PCR Positivity Rate in 17 Regional Health Departments in São Paulo in epidemiological weeks 10 to 35.

Figure 1 shows the positivity rate (T2) obtained by all RHD (Figure 1). The mean RHD score for the positivity rate was 0.256 between epidemiological weeks 10 to 35, and the variance was greater within an RHDs (0.126) than across RHDs (0.052). The most common score for the positivity rate indicator (T2) was 0 which indicates positivity rate greater than 19% (way above the minimum recommended by WHO (4)).

Three indicators were assigned zero values given that no data were publicly available: a) the diagnostic RT-PCR test turnaround time (T3); b) the repeated testing of individuals (T5); and, c) the percentage of cases interviewed for contact elicitation (T6).

Finally, every RHD with at least one local public lab received a score of one in the lab indicator (T4). In our sample, seventy percent of RHDs (12 RHDs) had at least one local public laboratory. Over the course of 25 weeks, the highest score of 2 was observed in 40.27% of the sample, an intermediate score of 1 was observed in 30.3%, and the lowest score in 29.41% of the sample. There was significant heterogeneity across the public labs in the processing capacity of RT-PCR tests per day. On September 17th, the range of variation of RT-PCR tests per day was between 0 (RHDs II, VII, and XI) and 4,500 (RHD I) (12).

Finally, the quality of RT-PCR testing data indicator was equal for all RHDs since there is only one database for all RHDs of the São Paulo State and this source reports the number of collected specimens.

**Figure 2.**
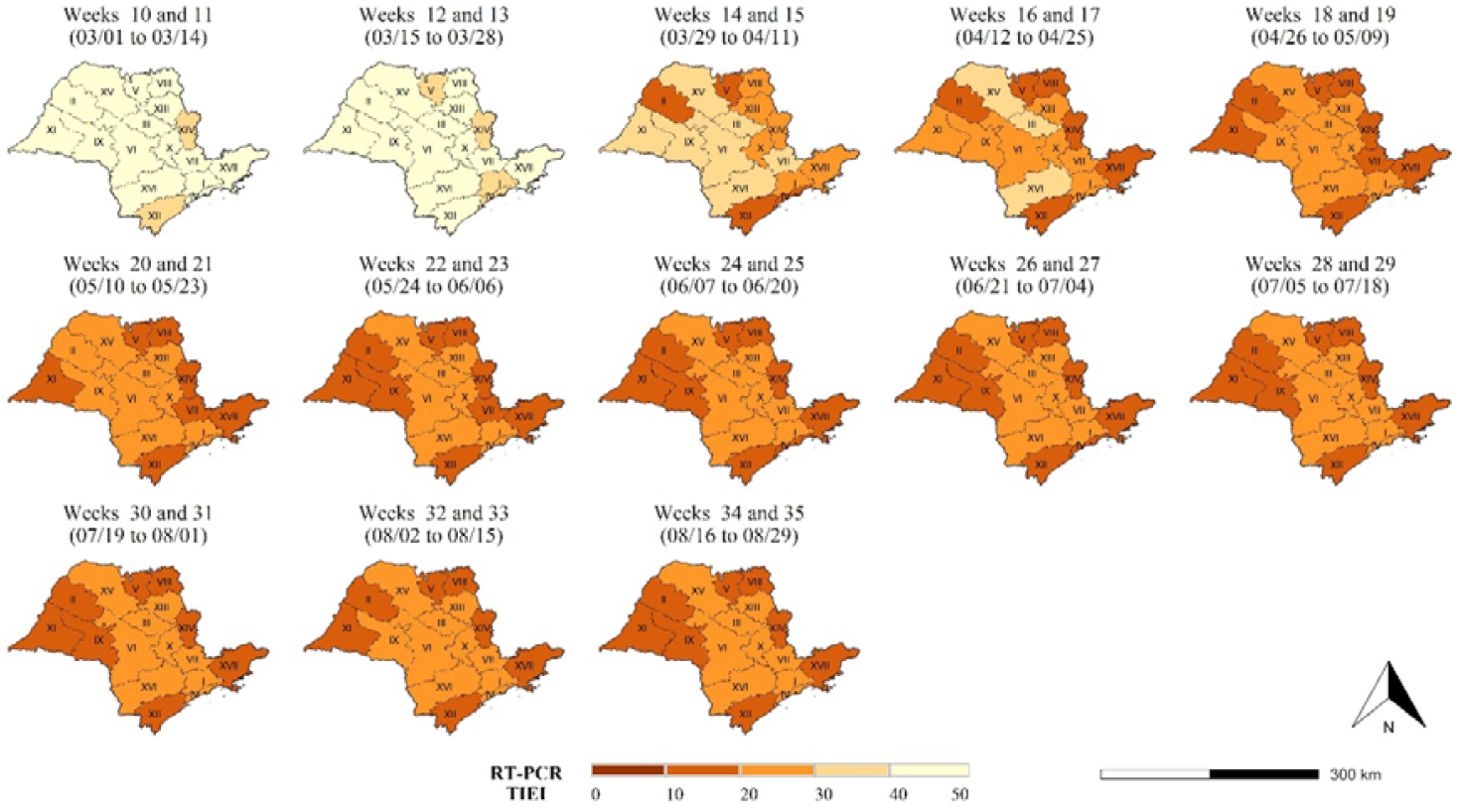
Index of Intensity Effort of RT-PCR Testing (RT-PCR TIEI) in 17 Regional Health Departments in São Paulo in epidemiological weeks 10 to 35.

Figure 2 shows the weekly score for the RT-PCR TIEI, where the darkest regions in the map have the lowest score. The maximum RT-PCR TIEI mean score of 42.35 was reached in week 11. The lowest mean of 15.88 was reached in week 25. The regions located in the northeast of São Paulo state and the coast consistently underperformed in their testing intensity efforts, whereas the central and western regions performed better. RHDs V, VIII, XII, XIV, and XVII had low scores since early on (epidemiological week 15). Overall, the RT-PCR TIEI in the final week of analysis is relatively low for all RHDs. The highest index score received was 20 (out of 100) in week 35: I, III, IV, VI, VII, X, XIII, XV, and XVI (n=9). The remaining RHDs scored 10 (n=5) and 15 (n=3).

**Figure 3.**
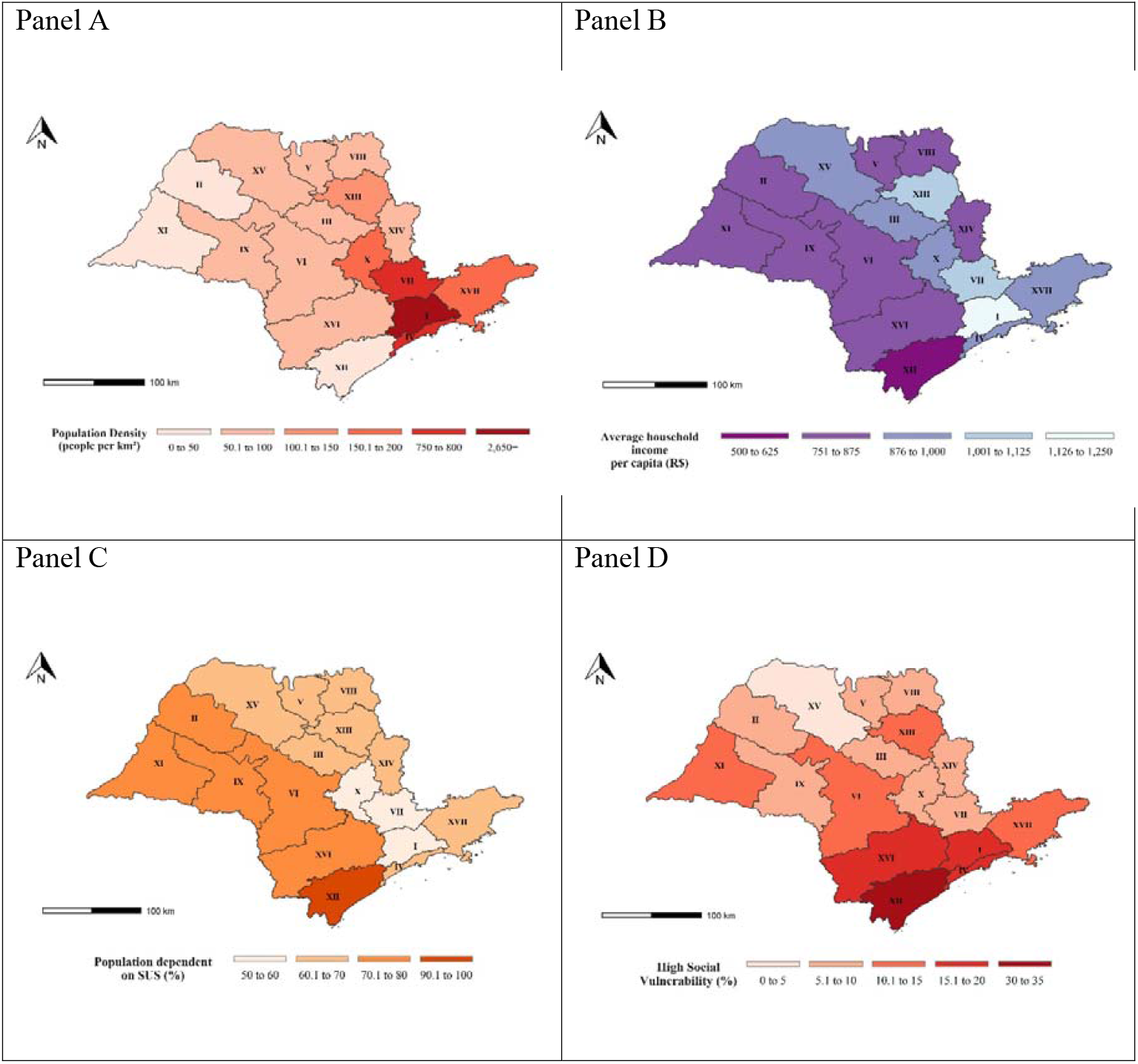
Population density (Panel A), Per Capita Income (Panel B), Proportion of People Dependent on SUS (Panel C), and Proportion of people living under high level of social vulnerability (Panel D) in 17 Regional Health Departments in São Paulo State.

Additionally, we analyzed how population density, income per capita, the proportion of the population dependent on SUS, and social vulnerability are related with the intensity of RT-PCR testing efforts in each region. Figure 3 reports the distribution of these variables in the 17 RHDs of São Paulo state. The regions with the highest population density in 2020 are RHD I, where the state’s capital is located, and RHDs IV e VII, all of them located in the central and coastal regions. However, the most vulnerable populations are located in the state’s coastal and southern regions, as Panels B, C, and D show. RHD XII presented the lowest values for per capita income, the proportion of population SUS dependent, and living under a high level of vulnerability.

Table 2 shows the results of the dynamic time-series cross-sectional models. We use four models to analyze how the RT-PCR TIEI is associated with each explanatory variable in the seventeen RHD between the epidemiological week 10 to 35. In the bottom panel of the table, we report the long-term predicted effects on the RT-PCR TIEI considering the lag dependent variable. In the long-run, the RT-PCR TIEI is negatively associated with socioeconomic vulnerability (p-value=0.000, 95% CI −0.887, −0.811), with a higher proportion of the population dependent on SUS (p-value= 0.000, 95% CI −0.871, −0.805) and with population density (p-value=0.000, 95% CI −0.853; −0.801). No evidence of first or second-order autocorrelation was found.

**Table 2.** Multiple regressions predicting association between RT-PCR TIEI and socioeconomic and demographic variables across RHDs.

|  | Model 1 |  | Model 2 |  | Model 3 |  | Model 4 |  |
| --- | --- | --- | --- | --- | --- | --- | --- | --- |
|  | beta | p-value [95% CI] | beta | p-value [95% CI] | beta | p-value [95% CI] | beta | p-value [95% CI] |
| Per Capita Income (R\$)<br>(2010) | 0.002 | 0.016* [0.000,0.003] | | | | | | |
| Population density<sub>2020</sub> |  |  | 0.000 | 0.238 [-0.000,0.000] |  |  |  |  |
| Share of Population SUS<br>Dependent<sub>2020</sub> |  |  |  |  | -0.012 | 0.377 [-0.039, 0.015] |  |  |
| High Social Vulnerability<br>(%)(<sub>2010</sub>) |  |  |  |  |  |  | -0.023 | 0.140 [-0.054, 0.008] |
| RT-PCR TIEI<sub>t-1</sub> | 0.823 | 0.000 ***[0.793, 0.853] | 0.827 | 0.000*** [0.799,0.856] | 0.826 | 0.000*** [0.798, 0.854] | 0.849 | 0.000*** [0.887, 0.811] |
| Constant | 1.195 | 0.058 [-0.048, 2.438] | 2.679 | 0.000*** [1.932, 3.426] | 3.533 | 0.003** [1.434, 5.631] | 3.016 | 0.000*** [2.122, 3.911] |
| Predicted Long-Term Effects: |  |  |  |  |  |  |  |  |
| Per Capita Income (R\$)(<sub>2010</sub>) | -0.821 | 0.000*** [-0.849,-0.792] | | | | | | |
| Population density<sub>2020</sub> |  |  | -0.827 | 0.000*** [-0.853, -0.801] |  |  |  |  |
| Share of Population SUS<br>Dependent<sub>2020</sub> |  |  |  |  | -0.838 | 0.000 [-0.871, -0.805] |  |  |
| High Social Vulnerability |  |  |  |  |  |  | -0.849 | 0.000 [-0.887, -0.811] |

|  | **Model 1** |  | **Model 2** |  | **Model 3** |  | **Model 4** |  |
| --- | --- | --- | --- | --- | --- | --- | --- | --- |
|  | **beta** | **p-value [95% CI]** | **beta** | **p-value [95% CI]** | **beta** | **p-value [95% CI]** | **beta** | **p-value [95% CI]** |
| (%)<sub>(2010)</sub> |  |  |  |  |  |  |  |  |
| Observations |  | 425 |  | 425 |  | 425 |  | 425 |
| Number of Weeks |  | 26 |  | 26 |  | 26 |  | 26 |
| RHDs |  | 17 |  | 17 |  | 17 |  | 17 |
| R-squared |  | 0.812 |  | 0.811 |  | 0.811 |  | 0.812 |
| Ar (1) |  | 0.126 |  | 0.132 |  | 0.131 |  | 0.129 |
| Ar (2) |  | 0.492 |  | 0.451 |  | 0.461 |  | 0.465 |

Notes: Standard errors clustered by RHD. The tests for autocorrelation in dynamic panel data (DPD) were implemented using Roodman (34) abar routine in Stata. Per capita income, population density, proportion of people living under high social vulnerability and dependency on the public health system (SUS) represent the mean for all municipalities belonging to each RHD.

## Discussion

The RT-PCR TIEI is a useful tool to analyze the intensity of efforts to control the SARS-CoV-2 pandemic and provide a metric to assess the public health testing efforts in association with other surveillance measures. Using this approach, our data show that between epidemiological week 10 to 35, the RT-PCR TIEI was highly heterogeneous across the 17 RHD in the state and trending down. The presence of a local public laboratory was the indicator with the highest impact on the final score. This finding is consistent with the lack of data for three indicators that received zero scores: (T3), (T5), and (T6). All RHDs presented a reduction in the RT-PCR TIEI over the 26-weeks analyzed. In addition, we found a negative association between the RT-PCR TIEI scores and the RHDs with the largest proportion of people living under high levels of social vulnerability, the population dependent on the public health system, and with higher population densities. In the course of the pandemic, the state of São Paulo advanced the intensity of its testing efforts in the public health system using RT-PCR tests for SARS-CoV-2 (12). However, the response was not homogenous and remained below WHO and CDC recommendations and the best practices adopted in other regions (Table 1). In consonance with our findings, the São Paulo State positivity rate averaged 30% on 18 October 2020, according to official data (12), which reveals that the state continues to rank below international counterparts, such as New York State (USA) (23).

Among the factors affecting the disparity in testing intensity across RHDs is the lack of a local laboratory, reflecting low past public health investments in infrastructure before the onset of the pandemic. The majority of RHDs have a local laboratory (12 of 17 RHDs). Laboratories are expensive and difficult to install during a public health emergency (24). Some regions were able to further increase their test processing capacity by partnering with universities and research institutes. However, the universities, laboratories, and research institutes collaborating in providing public access to RT-PCR are concentrated in only five RHDs, while others have public laboratories in their territories. There is no public laboratory in RHD XII (25), and its municipalities are among those with the lowest levels of social and economic development (15).

Indicator seven (the quality of RT-PCR testing data provided) permits examination of the ability to access information, which is fundamental to evaluate and monitor pandemic control measures (26). Since the beginning of the COVID-19 pandemic, the importance of sharing data has been underscored by the necessity to build and integrate computational infrastructure that can provide large datasets and facilitate data access by public servants, researchers, and the general population (26) (27). Although some RHDs claimed to have implemented contact tracing programs, such as RHDs I (28), III, and VI (29), an evaluation of contact-tracing efforts across all RHDs was not possible with currently available data.

Our results confirmed that socioeconomic vulnerability is a determining factor affecting access to RT-PCR testing and control strategies in São Paulo State. This finding is in line with Souza *et al* (30) that found an association between confirmed COVID-19 diagnosis and per capita income in São Paulo, the capital of São Paulo State. Given that demand for testing is not necessarily associated with regional testing supply, and further considering each region’s population density, our study’s findings highlight that structural inequities can contribute to the severity of the COVID-19 pandemic. Moreover, our findings provide evidence that underperformance in laboratory testing contributed to the emergency’s poor handling even in Brazil’s richest and most well-endowed state.

Differences in access to COVID-19 testing were also found in New York City by race and socioeconomic characteristics based on tested individuals’ ZIP codes (31). The city’s most vulnerable communities did not have proportional access to testing, and this disparity affected New York’s public health response. The same inequality related to COVID-19 testing access has been found in Brazil, where people living in areas with higher income per capita are more likely to have been tested (32), despite the difference between the health system in Brazil, which have a public health system, and the USA.

Considering the surveillance indicators, we identified three distinct COVID-19 testing scenarios. First, there were a few RHDs where the RT-PCR tests increased; in these cases, partnering with local universities and companies were instrumental. There was a second scenario in two RHDs where the laboratory capacity and the positivity rate were comparatively high. This is the case of RHD I and RHD IV, two of the most influential regions in the state since the onset of the pandemic in Brazil. In both regions, the number of private health care units and private laboratories may explain the higher number of cases and the number of RT-PCR tests performed in the public labs.

In the third scenario, which includes most of the RHDs in Sao Paulo, the number of tests performed was close to the number of cases recorded for most of the analyzed period. Most likely, RT-PCR testing was used in these RHDs to test only symptomatic hospitalized patients. Therefore, testing was not used to identify new cases and people who have been in contact with infected people.

There are limitations to this study. The Health Secretariat’s Intelligent Monitoring System does not report anonymized individual RT-PCR testing data conducted in private labs (12). Given these limitations, we were not able to estimate the diagnostic RT-PCR test turnaround time.

Given our goal of assessing RT-PCR testing efforts in public laboratories, our study also highlights that the Brazilian Health System (SUS) can play an instrumental role in reducing inequalities. Its decentralized structure allows for targeted policies, considering the distinct epidemiological profiles and vulnerabilities (33). During pandemics, control measures must be guided by the characteristics of the pandemic settings and the health system’s capacity in different locations. In this regard, the SUS allows municipalities and RHDs to adopt distinct strategies to test, identify, and isolate infected people, contact tracking, and quarantine infected people. Finally, our data suggest that in addition to the improvement in testing capacity, the autonomy of RHDs to modify their surveillance and control measures requires that federal and state authorities adopt policies that are aligned with these efforts, which was not the case through the course of the pandemic.

## Conclusion

The proposed RT-PCR testing intensity effort index allowed us to analyze the COVID-19 diagnostic testing policies in São Paulo State, the pandemic’s epicenter in Brazil. Since the obtained RT-PCR scores decline as the pandemic advanced, we conclude that the state’s testing intensity effort has been insufficient to control the SARS-CoV-2 transmission. The results confirm that socioeconomic vulnerable RHDs have lower RT-PCR TIEI scores, and local public laboratories are an important predictor of a higher RT-PCR TIEI score. Thus, the low RT-PCR TIEI and local laboratory capacity may affect the pandemic control, especially in the most socioeconomic vulnerable population.

## Data Availability

All data referred to in the manuscript are provided by open access platforms. The source of all data used in our manuscript are presented below:
Sao Paulo. Sistema de Monitoramento Inteligente - SIMI-SP [Internet]. Available: https://www.saopaulo.sp.gov.br/planosp/simi/
Sao Paulo State. Plano Sao Paulo [Internet]. Available: https://www.saopaulo.sp.gov.br/wp-content/uploads/2020/08/PlanoSP-apresentacao-v2.pdf
SEADE. Sistema de projecoes populacionais para os municipios do estado de Sao Paulo. [Internet]. Available:http://produtos.seade.gov.br/produtos/projpop/pdfs/projpop
Sao Paulo State. Indice Paulista de Vulnerabilidade Social [Internet]. Available: http://ipvs.seade.gov.br/view/pdf/ipvs/estado.pdf
Sao Paulo State. Estimativa da populacao SUS dependente (com base na saude suplementar). [Internet]. Available: http://tabnet.saude.sp.gov.br/deftohtm.exe?tabnet/ind47b_matriz.def
Brazilian Institute of Geography and Statistic (IBGE). Census 2010 [Internet]. Available: https://censo2010.ibge.gov.br/resultado

https://www.saopaulo.sp.gov.br/planosp/simi/testes/

http://tabnet.saude.sp.gov.br/deftohtm.exe?tabnet/ind47b_matriz.def

http://ipvs.seade.gov.br/view/pdf/ipvs/estado.pdf

https://censo2010.ibge.gov.br/resultado

http://produtos.seade.gov.br/produtos/projpop/pdfs/projpop

## Author Contributions

T.S., N.M., and L.B. conceived the research design, and completed the writing, review and editing of the manuscript. N.M., I.S., and M.Z. performed the statistical analyses. T.S., N.M., and L.B. have accessed and verified the underlying data. T.S., N.M., J.E.K, M.A.S.M.V, B.K. and L.B. interpreted the results and edited the final manuscript. All the authors discussed the results and contributed to the final manuscript.

## References

1. Reintjes R. Lessons in contact tracing from Germany. BMJ 2020;369:m2522. doi:10.1136/bmj.m2522 pmid:32586833.

2. Park YJ, Choe YJ, Park O, Park SY, Kim Y-M, Kim J, et al. Contact Tracing during Coronavirus Disease Outbreak, South Korea, 2020. Emerging Infect Dis. October, 2020;26(10):2465–8.

3. Centers for Disease Control and Prevention (CDC). SARS-CoV-2 Testing Strategy: Considerations for Non-Healthcare Workplaces. https://www.cdc.gov/coronavirus/2019-ncov/community/organizations/testing-non-healthcare-workplaces.html. Acessed September 23, 2020

4. World Health Organization. Public health criteria to adjust public health and social measures in the context of COVID-19. https://www.who.int/publications/i/item/public-health-criteria-to-adjust-public-health-and-social-measures-in-the-context-of-covid-19. Published May 12, 2020. Acessed August 31, 2020

5. World Health Organization. Laboratory testing strategy recommendations for COVID-19: interim guidance. https://www.who.int/publications/i/item/laboratory-testing-strategy-recommendations-for-covid-19-interim-guidance. Published March 21, 2020. Acessed August 31, 2020.

6. World Health Organization. COVID-19 Virtual Press Conferece. https://www.who.int/docs/default-source/coronaviruse/transcripts/who-audio-emergencies-coronavirus-press-conference-full-30mar2020.pdf. Published March 30, 2020. Acessed August 31, 2020

7. Mina, MJ, Parker, R, Larremore, DB. Rethinking Covid-19 Test Sensitivity - A Strategy for Containment. N Engl J Med. September 30, 2020.

8. Centers for Disease Control and Prevention (CDC). Evaluating Case Investigation and Contact Tracing Success website. https://www.cdc.gov/coronavirus/2019-ncov/php/contact-tracing/contact-tracing-plan/evaluating-success.html. Acessed September 23, 2020

9. Fiocruz, Instituto de Comunicação e Informação Científica e Tecnológica em Saúde (ICICT). MonitoraCovid-19 website. https://bigdata-covid19.icict.fiocruz.br/. Acessed August 31, 2020

10. São Paulo. Resolução SS No 43 DE 01/04/2020. Regulamenta, no âmbito do Estado de São Paulo, o fluxo para o diagnóstico do novo coronavírus - COVID-19, indica os laboratórios integrantes, e dá providências correlatas. https://www.legisweb.com.br/legislacao/?id=392206#:~:text=Regulamenta%2C%20no%20%C3%A2mbito%20do%20Estado,integrantes%2C%20e%20d%C3%A1%20provid%C3%AAncias%20correlatas.&text=III%20%2D%20Propiciar%2C%20atrav%C3%A9s%20do%20acervo,pesquisa%20no%20Instituto%20ou%20multic%C3%AAntricos. Published April 4, 2020. Acessed August 31, 2020

11. Candido DS, Claro IM, Jesus JG, Souza WM, Moreira FRR, Dellicour S, et al. Evolution and epidemic spread of SARS-CoV-2 in Brazil. MedRxiv 2020; 23 jun. https://www.medrxiv.org/content/10.1101/2020.06.11.20128249v2

12. São Paulo. Sistema de Monitoramento Inteligente - SIMI-SP website. https://www.saopaulo.sp.gov.br/planosp/simi/. Acessed August 31, 2020.

13. São Paulo State. Plano São Paulo.https://www.saopaulo.sp.gov.br/wp-content/uploads/2020/08/PlanoSP-apresentacao-v2.pdf. Acessed August 31, 2020

14. SEADE. Sistema de projeções populacionais para os municípios do estado de São Paulo website. http://produtos.seade.gov.br/produtos/projpop/pdfs/projpop. Acessed August 31, 2020

15. São Paulo State. Índice Paulista de Vulnerabilidade Social. http://ipvs.seade.gov.br/view/pdf/ipvs/estado.pdf. xPublished 2010. Acessed August 31, 2020

16. São Paulo State. Estimativa da população SUS dependente (com base na saúde suplementar) website. http://tabnet.saude.sp.gov.br/deftohtm.exe?tabnet/ind47b_matriz.def. Acessed August 31, 2020

17. Brazilian Institute of Geography and Statistics (IBGE). Census 2010 website. https://censo2010.ibge.gov.br/resultados.html. Acessed August 31, 2020.

18. Brown School of Public Health, Microsoft AI for Health, Harvard Global Health Institute. New Testing Targets: As COVID-19 outbreaks grow more severe, most U.S. states still fall far short on testing website. https://globalepidemics.org/july-6-2020-state-testing-targets/. Acessed October 26, 2020

19. The Atlantic. The COVID Tracking Project: Counting COVID-19 Tests: How States Do It, How We Do It, and What’s Changing website. https://covidtracking.com/blog/counting-covid-19-tests. Acessed October 26, 2020.

20. Larremore DB, Wilder B, Lester E, et al. Test sensitivity is secondary to frequency and turnaround time for COVID-19 surveillance. Preprint. medRxiv. 2020;2020.06.22.20136309. Published 2020 Jun 27. doi:10.1101/2020.06.22.20136309

21. Johns Hopkins University. COVID-19 Dashboard by the Center for Systems Science and Engineering (CSSE) website. Coronavirus Resource Center. https://coronavirus.jhu.edu/map.html. Acessed October 26, 2020.

22. De Boef, Suzanna, and Luke Keele. “Taking Time Seriously.” American Journal of Political Science 52, no. 1 (2008): 184–200. Accessed October 29, 2020. http://www.jstor.org/stable/25193805.

23. New York State. NYS-COVID19-Tracker website https://covid19tracker.health.ny.gov/views/NYS-COVID19-Tracker/NYSDOHCOVID-19Tracker-ap?%3Aembed=yes&%3Atoolbar=no&%3Atabs=n. Acessed August 31, 2020

24. Nhan C, Laprise R, Douville-Fradet M, Macdonald ME, Quach C. Coordination and resource-related difficulties encountered by Quebec’s public health specialists and infectious diseases/medical microbiologists in the management of A (H1N1) - a mixed-method, exploratory survey. BMC Public Health. Feb 10, 2012;12(1):115.

25. São Paulo State. Laboratórios Regionais - Instituto Adolfo Lutz website. http://www.ial.sp.gov.br/ial/lab.-regionais/. Acessed September 23, 2020.

26. Xu B, Kraemer MUG, Xu B, Gutierrez B, Mekaru S, Sewalk K, et al. Open access epidemiological data from the COVID-19 outbreak. The Lancet Infectious Diseases. May 2020;20(5):534.

27. Heymann DL. Data sharing and outbreaks: best practice exemplified. The Lancet. Feb 2020;395(10223):469–70.

28. Leal MC, Muniz LF, Ferreira TS, Santos CM, Almeida LC, Van Der Linden V, et al. Hearing Loss in Infants with Microcephaly and Evidence of Congenital Zika Virus Infection - Brazil, November 2015-May 2016. MMWR Morbidity and mortality weekly report. September 2016;65(34):917–9.

29. G1 São Carlos e Araraquara. Araraquara é uma das 3 cidades escolhidas para projeto piloto de rastreamento da Covid-19.https://g1.globo.com/sp/sao-carlos-regiao/noticia/2020/07/09/araraquara-e-uma-das-3-cidades-escolhidas-para-projeto-piloto-de-rastreamento-da-covid-19.ghtml. Published July 9, 2020. Acessed September 23, 2020

30. de Souza, W.M., Buss, L.F., Candido, D.d.S. et al. Epidemiological and clinical characteristics of the COVID-19 epidemic in Brazil. Nat Hum Behav 4, 856–865 (2020). https://doi.org/10.1038/s41562-020-0928-4

31. Lieberman-Cribbin W, Tuminello S, Flores RM, Taioli E. Disparities in COVID-19 Testing and Positivity in New York City. American Journal of Preventive Medicine. September, 2020;59(3):326–32.

32. Brazilian Institute of Geography and Statistic (IBGE). Pesquisa Nacional por Amostra de DomicíliosLJ: PNAD COVID19LJ: resultado mensal / IBGE, Coordenação de Trabalho e Rendimento website https://biblioteca.ibge.gov.br/index.php/biblioteca-catalogo?view=detalhes&id=2101755. Published August, 2020. Acessed September 23, 2020

33. Jaccoud L, Vieira FS. Federalismo, Integralidade e Autonomia no SUS: Desvinculação da aplicação de recursos federais e os desafios da coordenação. 2018 https://www.ipea.gov.br/portal/images/stories/PDFs/TDs/td_2399.pdf

34. Roodman, D. How to do xtabond2: An introduction to difference and system GMM in Stata. The Stata Journal. 2009 Mar; 9 (1): 86–136.

