## Appendix 1 for "Using RT-PCR Testing to Assess the Effectiveness of Outbreak Control Efforts in São Paulo State, the Pandemic’s Epicenter in Brazil, according to Socioeconomic Vulnerabilities"

| **Appendix 1. Municipalities, Number of Local Public Laboratory RT-PCR, Total and Percentage of Population of each of the 17 Regional Health Departments of the São Paulo State in August 2020.** | | | | | |
| --- | --- | --- | --- | --- | --- |
| **RHD code** | **RHD Name** | **Municipalities** | **Number of Local Laboratories** | **Population (%)** | **Population dependent on the Public Health System (%)** |
| **RHD 1** | **Grande São Paulo** | **Arujá, Barueri, Biritiba-Mirim, Caieiras, Cajamar, Carapicuíba, Cotia, Diadema, Embu, Embu-Guaçu, Ferraz de Vasconcelos, Francisco Morato, Franco da Rocha, Guararema, Guarulhos, Itapecerica da Serra, Itapevi, Itaquaquecetuba, Jandira, Juquitiba, Mairiporã, Mauá, Mogi das Cruzes, Osasco, Pirapora do Bom Jesus, Poá, Ribeirão Pires, Rio Grande da Serra, Salesópolis, Santa Isabel, Santana de Parnaíba, Santo André, São Bernardo do Campo, São Caetano do Sul, São Lourenço da Serra, São Paulo, Suzano, Taboão da Serra, Vargem Grande Paulista.** | **4** | **21138247 (47.35%)** | **56%** |
| **RHD 2** | **Araçatuba** | **Alto Alegre, Andradina, Araçatuba, Auriflama, Avanhandava, Barbosa, Bento de Abreu, Bilac, Birigui, Braúna, Brejo Alegre, Buritama, Castilho, Clementina, Coroados, Gabriel Monteiro, Glicério, Guaraçaí, Guararapes, Guzolândia, Ilha Solteira, Itapura, Lavínia, Lourdes, Luiziânia, Mirandópolis, Murutinga Do Sul, Nova Castilho, Nova Independência, Nova Luzitânia, Penápolis, Pereira Barreto, Piacatu, Rubiácea, Santo Antônio Do Aracanguá, Santópolis do Aguapeí, Sud Mennucci, Suzanápolis, Turiúba, Valparaíso.** | **1** | **764041 (1.71%)** | **76%** |
| **RHD 3** | **Araraquara** | **Américo Brasiliense, Araraquara, Boa Esperança do Sul, Borborema, Cândido Rodrigues, Descalvado, Dobrada, Dourado, Gavião Peixoto, Ibaté, Ibitinga, Itápolis, Matão, Motuca, Nova Europa, Porto Ferreira, Ribeirão Bonito, Rincão, Santa Ernestina, Santa Lúcia, São Carlos, Tabatinga, Taquaritinga, Trabiju.** | **1** | **991435 (2.22%)** | **63%** |
| **RHD 4** | **Baixada Santista** | **Bertioga, Cubatão, Guarujá, Itanhaém, Mongaguá, Peruíbe, Praia Grande, Santos, São Vicente.** | **1** | **1831884 (4.1%)** | **63%** |
| **RHD 5** | **Barretos** | **Altair, Barretos, Bebedouro, Cajobi, Colina, Colômbia, Guaíra, Guaraci, Jaborandi, Monte Azul Paulista, Olímpia, Severínia, Taiaçu, Taiúva, Taquaral, Terra Roxa, Viradouro, Vista Alegre do Alto.** | **0** | **425090 (0.95%)** | **63%** |
| **RHD 6** | **Bauru** | **Águas De Santa Bárbara, Agudos, Anhembi, Arandu, Arealva, Areiópolis, Avaí, Avaré, Balbinos, Barão De Antonina, Bariri, Barra Bonita, Bauru, Bocaina, Bofete, Boracéia, Borebi, Botucatu, Brotas, Cabrália Paulista, Cafelândia, Cerqueira César, Conchas, Coronel Macedo, Dois Córregos, Duartina, Fartura, Getulina, Guaiçara, Iacanga, Iaras, Igaraçu do Tietê, Itaí, Itaju, Itaporanga, Itapuí, Itatinga, Jaú, Laranjal Paulista, Lençóis, Paulista, Lins, Lucianópolis, Macatuba, Manduri, Mineiros Do Tietê, Paranapanema, Pardinho, Paulistânia, Pederneiras, Pereiras, Piraju, Pirajuí, Piratininga, Pongaí, Porangaba, Pratânia, Presidente Alves, Promissão, Reginópolis, Sabino, São Manuel, Sarutaiá, Taguaí, Taquarituba, Tejupá, Torre de Pedra, Torrinha, Uru.** | **2** | **1741281 (3.9%)** | **76%** |
| **RHD 7** | **Campinas** | **Águas De Lindóia, Americana, Amparo, Artur Nogueira, Atibaia, Bom Jesus Dos Perdões, Bragança Paulista, Cabreúva, Campinas, Campo Limpo Paulista, Cosmópolis, Holambra, Hortolândia, Indaiatuba, Itatiba, Itupeva, Jaguariúna, Jarinu, Joanópolis, Jundiaí, Lindóia, Louveira, Monte Alegre do Sul, Monte Mor, Morungaba, Nazaré Paulista, Nova Odessa, Paulínia, Pedra Bela, Pedreira, Pinhalzinho, Piracaia, Santa Bárbara D'Oeste, Santo Antônio da Posse, Serra Negra, Socorro, Sumaré, Tuiuti, Valinhos, Vargem, Várzea Paulista, Vinhedo.** | **2** | **4562125 (10.21%)** | **57%** |
| **RHD 8** | **Franca** | **Aramina, Buritizal, Cristais Paulista, Franca, Guará, Igarapava, Ipuã, Itirapuã, Ituverava, Jeriquara, Miguelópolis, Morro Agudo, Nuporanga, Orlândia, Patrocínio Paulista, Pedregulho, Restinga, Ribeirão Corrente, Rifaina, Sales Oliveira, São Joaquim da Barra, São José da Bela Vista.** | **0** | **696336 (1.56%)** | **67%** |
| **DRS 9** | **Marília** | **Adamantina, Álvaro de Carvalho, Alvinlândia, Arco Íris, Assis, Bastos, Bernardino de Campos, Borá, Campos Novos Paulista, Cândido Mota, Canitar, Chavantes, Cruzália, Echaporã, Espírito Santo do Turvo, Fernão, Flórida Paulista, Florínia, Gália, Garça, Guaimbê, Guarantã, Herculândia, Iacri, Ibirarema, Inúbia Paulista, Ipaussu, Júlio Mesquita, Lucélia, Lupércio, Lutécia, Maracaí, Mariápolis, Marília, Ocauçu, Óleo, Oriente, Oscar Bressane, Osvaldo Cruz, Ourinhos, Pacaembu, Palmital, Paraguaçu Paulista, Parapuã, Pedrinhas Paulista, Platina, Pompéia, Pracinha, Queiroz, Quintana, Ribeirão do Sul, Rinópolis, Sagres, Salmourão, Salto Grande, Santa Cruz do Rio Pardo, São Pedro do Turvo, Tarumã, Timburi, Tupã, Ubirajara, Vera Cruz.** | **1** | **1109670 (2.5%)** | **79%** |
| **DRS 10** | **Piracicaba** | **Águas de São Pedro, Analândia, Araras, Capivari, Charqueada, Conchal, Cordeirópolis, Corumbataí, Elias Fausto, Engenheiro Coelho, Ipeúna**  **Iracemápolis, Itirapina, Leme, Limeira, Mombuca, Piracicaba, Pirassununga, Rafard, Rio Claro, Rio das Pedras, Saltinho, Santa Cruz da Conceição, Santa Gertrudes, Santa Maria da Serra, São Pedro.** | **2** | **1539600 (3.45%)** | **57%** |
| **DRS 11** | **Presidente Prudente** | **Alfredo Marcondes, Álvares Machado, Anhumas, Caiabu, Caiuá, Dracena, Emilianópolis, Estrela do Norte, Euclides da Cunha Paulista, Flora Rica, Iepê, Indiana, Irapuru, João Ramalho, Junqueirópolis, Marabá Paulista, Martinópolis, Mirante do Paranapanema, Monte Castelo, Nantes, Narandiba, Nova Guataporanga, Ouro Verde, Panorama, Paulicéia, Piquerobi, Pirapozinho, Presidente Bernardes, Presidente Epitácio, Presidente Prudente, Presidente Venceslau, Quatá, Rancharia, Regente Feijó, Ribeirão dos Índios, Rosana, Sandovalina, Santa Mercedes, Santo Anastácio, Santo Expedito, São João do Pau D'Alho, Taciba, Tarabai, Teodoro Sampaio, Tupi Paulista.** | **1** | **752260 (1.68%)** | **77%** |
| **DRS 12** | **Registro** | **Barra do Turvo, Cajati, Cananéia, Eldorado, Iguape, Ilha Comprida, Iporanga, Itariri, Jacupiranga, Juquiá, Miracatu, Pariquera-Açu, Pedro de Toledo, Registro, Sete Barras.** | **0** | **278754 (0.62%)** | **91%** |
| **DRS 13** | **Ribeirão Preto** | **Altinópolis, Barrinha, Batatais, Brodowski, Cajuru, Cássia dos Coqueiros, Cravinhos, Dumont, Guariba, Guatapará, Jaboticabal, Jardinópolis, Luís Antônio, Monte Alto, Pitangueiras, Pontal, Pradópolis, Ribeirão Preto, Santa Cruz da Esperança, Santa Rita do Passa Quatro, Santa Rosa de Viterbo, Santo Antônio da Alegria, São Simão, Serra Azul, Serrana, Sertãozinho.** | **3** | **1477530 (3.3%)** | **62%** |
| **DRS 14** | **São João da Boa Vista** | **Aguaí, Águas da Prata, Caconde, Casa Branca, Divinolândia, Espírito Santo do Pinhal, Estiva Gerbi, Itapira, Itobi, Mococa, Mogi Guaçu, Mogi Mirim, Santa Cruz das Palmeiras, Santo Antônio do Jardim, São João da Boa Vista, São José do Rio Pardo, São Sebastião da Grama, Tambaú, Tapiratiba, Vargem Grande do Sul.** | **0** | **809836 (1.81%)** | **69%** |
| **DRS 15** | **São José do Rio Preto** | **Adolfo, Álvares Florence, Américo de Campos, Aparecida D'Oeste, Ariranha, Aspásia, Bady Bassitt, Bálsamo, Cardoso, Catanduva, Catiguá, Cedral, Cosmorama, Dirce Reis, Dolcinópolis, Elisiário, Embaúba, Estrela D'Oeste, Fernandópolis, Fernando Prestes, Floreal, Gastão Vidigal, General, Salgado, Guapiaçu, Guarani D'Oeste, Ibirá, Icém, Indiaporã, Ipiguá, Irapuã, Itajobi, Jaci, Jales, José Bonifácio, Macaubal, Macedônia, Magda, Marapoama, Marinópolis, Mendonça, Meridiano, Mesópolis, Mira Estrela, Mirassol, Mirassolândia, Monções, Monte Aprazível, Neves Paulista, Nhandeara, Nipoã, Nova Aliança, Nova Canaã Paulista, Nova Granada, Novais, Novo Horizonte, Onda Verde, Orindiúva, Ouroeste, Palestina, Palmares Paulista, Palmeira D'Oeste, Paraíso, Paranapuã, Parisi, Paulo de Faria, Pedranópolis, Pindorama, Pirangi, Planalto, Poloni, Pontalinda, Pontes Gestal, Populina, Potirendaba, Riolândia, Rubinéia, Sales, Santa Adélia, Santa Albertina, Santa Clara D'Oeste, Santa Fé do Sul, Santa Rita D'Oeste, Santa Salete, Santana da Ponte Pensa, São Francisco, São João das Duas Pontes, São João de Iracema, São José do Rio Preto, Sebastianópolis do Sul, Tabapuã, Tanabi, Três Fronteiras, Turmalina, Ubarana, Uchoa, União Paulista, Urânia, Urupês, Valentim Gentil, Vitória Brasil, Votuporanga, Zacarias.** | **2** | **1570421 (3.51%)** | **68%** |
| **DRS 16** | **Sorocaba** | **Alambari, Alumínio, Angatuba, Apiaí, Araçariguama, Araçoiaba da Serra, Barra do Chapéu, Boituva, Bom Sucesso de Itararé, Buri, Campina do Monte Alegre, Capão Bonito, Capela do Alto, Cerquilho, Cesário Lange, Guapiara, Guareí, Ibiúna, Iperó, Itaberá, Itaóca, Itapetininga, Itapeva, Itapirapuã Paulista, Itararé, Itu, Jumirim, Mairinque, Nova Campina, Piedade, Pilar do Sul, Porto Feliz, Quadra, Ribeira, Ribeirão Branco, Ribeirão Grande, Riversul, Salto, Salto de Pirapora, São Miguel Arcanjo, São Roque, Sarapuí, Sorocaba, Tapiraí, Taquarivaí, Tatuí, Tietê, Votorantim.** | **1** | **2461760 (5.51%)** | **72%** |
| **DRS 17** | **Taubaté** | **Aparecida, Arapeí, Areias, Bananal, Caçapava, Cachoeira Paulista, Campos do Jordão, Canas, Caraguatatuba, Cruzeiro, Cunha, Guaratinguetá, Igaratá, Ilhabela, Jacareí, Jambeiro, Lagoinha, Lavrinhas, Lorena, Monteiro Lobato, Natividade da Serra, Paraibuna, Pindamonhangaba, Piquete, Potim, Queluz, Redenção da Serra, Roseira, Santa Branca, Santo Antônio do Pinhal, São Bento do Sapucaí, São José do Barreiro, São José dos Campos, São Luiz do Paraitinga, São Sebastião, Silveiras, Taubaté, Tremembé, Ubatuba.** | **0** | **2489629 (5.58%)** | **69%** |

**Source: (8) (11) (30).**
